# Long-Term Seizure Reduction Associated with Vagal Nerve Stimulation in Dravet Syndrome

**DOI:** 10.1101/2024.12.30.24319582

**Authors:** Sunanjay Bajaj, Alina Ivaniuk, Tobias Bruenger, Émile Moura Coelho Da Silva, Emily Huth, Ludovica Montanucci, Costin Leu, Gary Taylor, Mousumi Sinha, Rahil A. Tai, Manish N. Shah, Michael W. Watkins, Jeremy E. Lankford, Indira M. Kommuru, Sandipan Pati, Prakash Kotagal, Andreas Alexopoulos, Samden D. Lhatoo, Elia Pestana Knight, Gretchen Von Allmen, Dennis Lal

## Abstract

*SCN1A* variants cause a range of epilepsy syndromes, including Dravet syndrome, leading to early cognitive and functional impairment. Despite advances in medical management, drug-resistant epilepsy remains common. Vagal nerve stimulation (VNS) has been suggested reducing seizure frequency in these patients but there is a lack of long-term follow-up, quantitative analysis that corrected for confounding factors such as antiseizure medications (ASMs) and the impact of VNS settings on response.

This two-center, retrospective cohort study analyzed 12-month and for the first time up to ten-year seizure outcomes in therapy-refractory epilepsy patients with loss-of-function *SCN1A* variants (93.75% Dravet Syndrome) who underwent VNS implantation. A ≥50% seizure frequency reduction was observed in 93.75% (15/16) of patients in the 12-month and 87.5% (14/15) in the ten-year period. Median seizure frequency was significantly lower in both follow-up periods than in the pre-implantation period. Linear mixed-effects regression showed that the reduction in seizure burden was independent of ASM use, and the VNS duty cycle was significantly associated with seizure reduction. Three individuals (18.8%) experienced minor side effects.

Our results highlight the benefits of genotype-driven therapeutic interventions such as VNS in patients with *SCN1A*-related epilepsy. This study emphasizes the need for further implementation of genotype-driven clinical decision-making.

## Introduction

Variants in *SCN1A*, encoding the alpha subunit of the voltage-gated sodium channel Na_V_1.1, are associated with several epilepsy syndromes, including two types of epileptic encephalopathies and Generalized Epilepsy with Febrile Seizures Plus (GEFS+). Dravet Syndrome (DS) is the most-well studied among these disorders and is characterized by early-onset drug-resistant epilepsy (DRE), severe developmental delays, and cognitive impairments^1^. The early onset of seizures (<12 months) and resistance to treatment often result in significant morbidity and reduction in the overall quality of life for patients and their families^1^.

Given the early drug resistance in *SCN1A*-related epilepsy, the use of neuromodulation strategies such as Vagal Nerve Stimulation (VNS) is increasing^2,3^. Multiple studies have shown the benefit of VNS in achieving seizure frequency reduction in DS patients^4,5^. However, the proportion of responders – defined as patients with ≥50% seizure reduction – has been variable (33-75%)^4,5^. Moreover, the long-term efficacy of VNS for seizure frequency reduction is unclear as post-VNS follow-up periods have been short (12-36 months)^4,5^. In addition, there is a knowledge gap regarding potential confounding effects of anti-seizure medications (ASM) and whether certain VNS settings affect response^6^.

To address these gaps, we aimed to define the long-term efficacy of VNS in reducing seizure frequency and quantify the impact of ASM and VNS settings on the observed benefit in patients with DS. We assessed both short-term (12 months) and long-term (up to ten years) outcomes after VNS implantation. Overall, we observed long-term efficacy of VNS while controlling for confounding factors and identified VNS settings associated with response.

## Material and Methods

### Patient Cohort Selection

Patients were retrospectively identified over a 21-year period (04/2003 – 10/2024) from the databases of tertiary epilepsy centers at UTHealth Houston/Memorial Hermann Hospital (MHH) and Cleveland Clinic. The following criteria were used: (1) presence of pathogenic or likely pathogenic *SCN1A* missense or truncating variants per American College of Medical Genetics (ACMG) criteria^7^, (2) diagnosis of DRE, defined as seizure persistence despite treatment with ≥two adequately dosed ASM, and (3) ≥12 months of post-VNS follow-up. Sixteen patients met these criteria, with 14 from UTHealth and two from Cleveland Clinic.

Epilepsy phenotypes were categorized as DS or GEFS+ per International League Against Epilepsy (ILAE) criteria^1,8,9^. Of these 16 patients, 93.75% (15/16) were diagnosed with DS based on their presentation with febrile and afebrile focal or generalized tonic-clonic seizures within the first year of life with no prior history of developmental concerns^9,10^. One patient presented with late-onset seizures (24 months) but refractory dialeptic seizures requiring VNS, and the development was normal; thus, the patient was categorized as having GEFS +^1^.

### Clinical Data Collection and Curation

Demographic, seizure, ASM, and comorbidity data were extracted from longitudinal electronic health records (EHRs) at UTHealth/MHH, Cleveland Clinic and prior EHR available via data sharing (Care Everywhere; https://www.epic.com/careeverywhere/). Seizure semiology was classified per the ILAE glossary^11^. Age at seizure onset was calculated from birth to first recorded seizure, and seizure frequency standardized as monthly clinical seizure frequency (MCSF) in 28-day intervals. The pre-VNS period covered 12 months before VNS, with follow-up divided into short-term (12 months) and long-term (up to ten years). Responders were defined as those achieving ≥50% seizure reduction post-VNS.

### Variant Pathogenicity Classification

The cohort included 15 patients with single nucleotide variants (SNVs) and one exon deletion in *SCN1A* identified through clinical genetic testing (Table S1). Variant pathogenicity (re)classification followed ACMG criteria. We followed the recent variant curation expert panel (VCEP) guidelines developed for voltage-gaged sodium channels (https://clinicalgenome.org/docs/clingen-variant-curation-expert-panel-vcep-protocol/). Further resources used were the SCN gene portal (https://scn-viewer.broadinstitute.org/; https://scn1a-prediction-model.broadinstitute.org), SpliceAI (v1.3.1; https://github.com/Illumina/SpliceAI/) for splice variant assessments, gnomAD (v4.0.1; https://gnomad.broadinstitute.org/) for population frequency assessments.

### Variant Visualization

Pymol (v.3.1; https://www.pymol.org/) was used for 3D protein structure visualization (Figure 1A).

**Figure 1.**
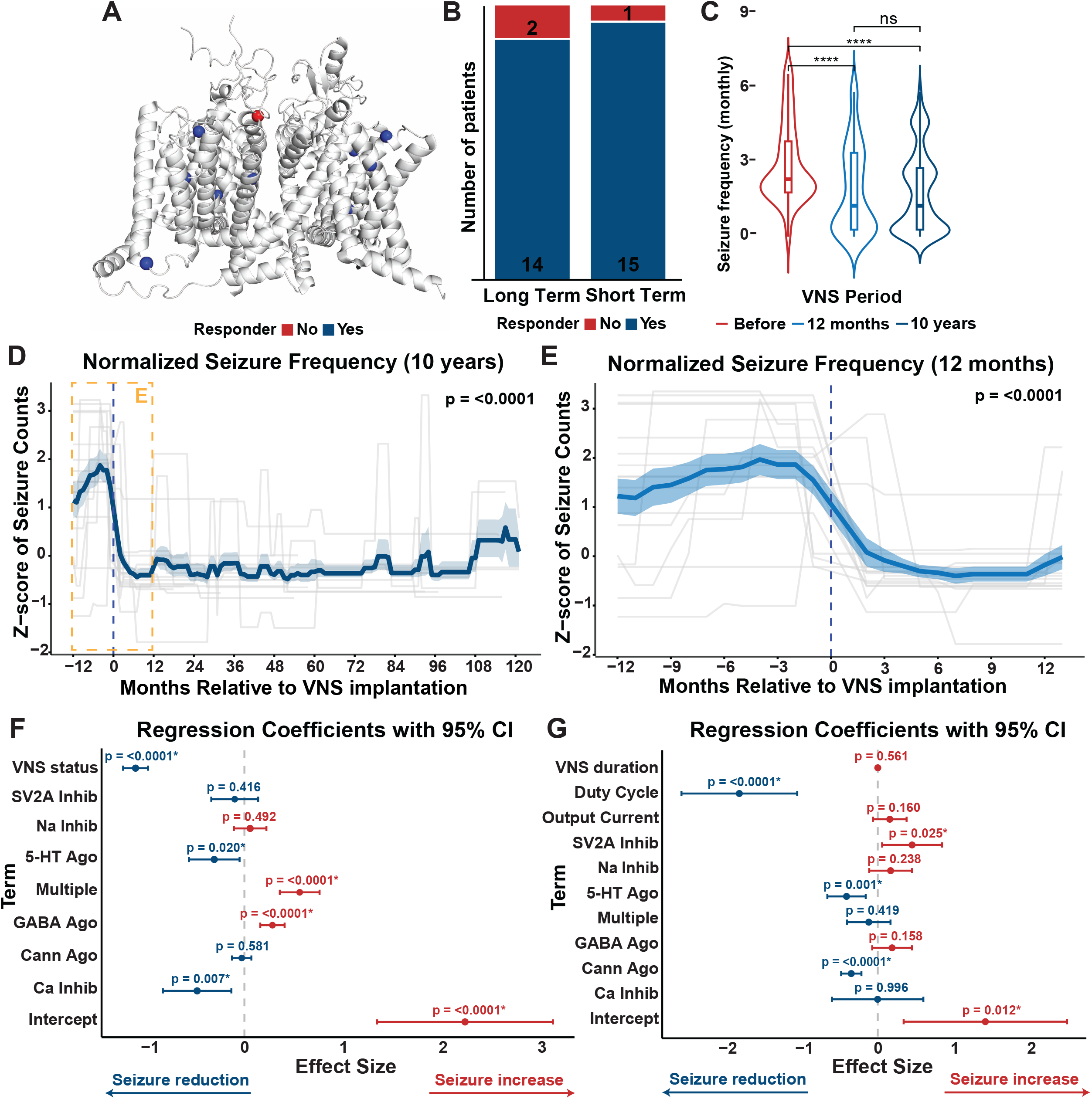
A. 3D structure showing missense variants in the cohort. Blue and red dots represent the variant sites of the responders and non-responders, respectively. B. Bar plots showing short-term (12 months) and long-term (10 years) VNS response. 15/16 (93.8%) of patients responded to VNS in the short term and 14/16 (87.5%) responded in the long term. Response defined as ≥50% reduction in median seizure frequency. Black: absolute counts C. Violin plots showing cross-sectional change in seizure frequency (logarithm of absolute counts) before and after VNS implantation. There is a significant reduction in seizure frequency in the short-term (12 months) and long-term period (10 years). Wilcoxon Sum Test used for significance: * <0.05, ** <0.01, *** <0.001, **** <0.0001. D. Normalized seizure burden (Z-score of absolute seizure counts) at 10-year follow-up. There is a significant reduction in mean seizure frequency (dark blue line) after VNS implantation (dashed blue line). Seizure frequencies of individual patients are visualized using the grey lines. Significance calculated using Wilcoxon Sum Test. Yellow box indicates period shown in panel E. E. Normalized seizure burden (Z-score of absolute seizure counts) at 12-month follow-up. There is a significant reduction in mean seizure frequency (light blue line) after VNS implantation (dashed blue line). Seizure frequencies of individual patients are visualized using the grey lines. Significance calculated using Wilcoxon Sum Test. F. Forest plot showing estimated coefficients (dots) with 95% confidence intervals (bars) from a linear mixed-effects regression model analyzing effects of VNS status (binary variable indicating status of VNS implantation (Y/N)) and ASM on the seizure frequency. Negative estimates indicate reduction, and positive estimators indicate increase in seizure frequency. VNS implantation, calcium channel inhibitors and serotonin agonists are associated with a significant reduction while ASM targeting multiple receptors and GABA agonists are associated with an increase in seizure frequency. G. Forest plot showing estimated coefficients (dots) with 95% confidence intervals (bars) from a linear mixed-effects regression model analyzing effects of duration of VNS duration, VNS duty cycle and generator current, and ASM on seizure frequency. VNS duty cycle, serotonin agonists, and cannabinoid agonists are associated with a significant reduction, while SV2A inhibitors are associated with an increase in seizure frequency.

### Statistical Analysis

All analyses were performed in R (v2024.0.2). Normality was assessed using the Kolmogorov-Smirnov test (In-built R function), with medians and quartiles summarizing cohort characteristics (Table 1). A two-month VNS ramp-up period was excluded. Seizure frequency was compared using the Wilcoxon signed-rank test. Longitudinal analysis involved Z-score normalization of seizure frequency to baseline, calculated as Z = (X – μ)/ σ, where X = seizure count, μ = mean seizure count, and σ = standard deviation. Paired differences were analyzed using the Wilcoxon signed-rank test. Linear mixed-effects regression models assessed VNS and ASM effect sizes on seizure frequency (log-normalized), adjusting for random (patient-specific) effects and testing multicollinearity among fixed effects.

**Table 1.**
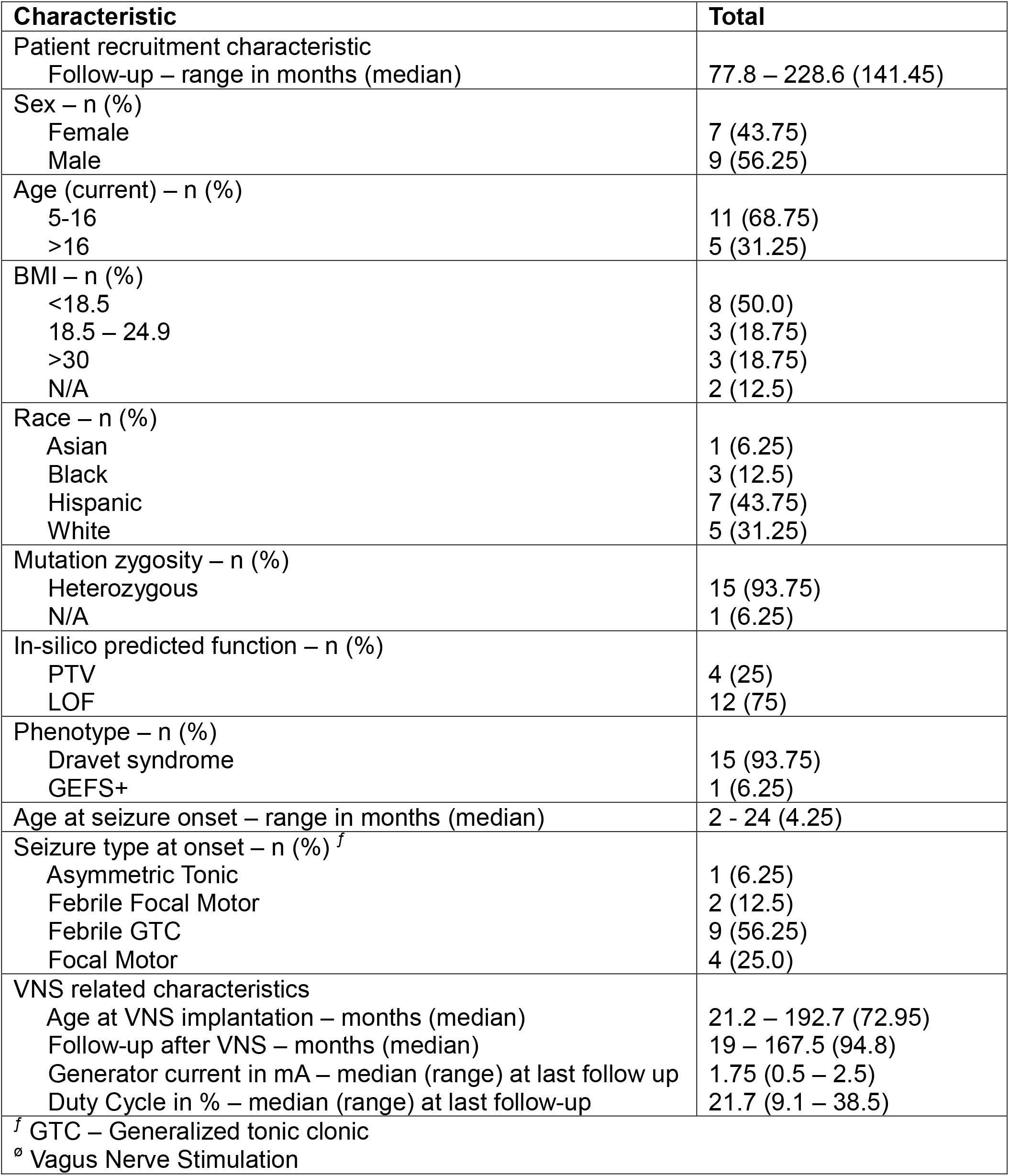
Patient Characteristics. Baseline characteristics of this cohort of 16 patients. Units for each value are specified alongside the respective characteristic.

| **Characteristic** | **Total** |
| --- | --- |
| Patient recruitment characteristic<br>Follow-up – range in months (median) | 77.8 – 228.6 (141.45) |
| Sex – n (%)<br>Female<br>Male | 7 (43.75)<br>9 (56.25) |
| Age (current) – n (%)<br>5-16<br>>16 | 11 (68.75)<br>5 (31.25) |
| BMI – n (%)<br><18.5<br>18.5 – 24.9<br>>30<br>N/A | 8 (50.0)<br>3 (18.75)<br>3 (18.75)<br>2 (12.5) |
| Race – n (%)<br>Asian<br>Black<br>Hispanic<br>White | 1 (6.25)<br>3 (12.5)<br>7 (43.75)<br>5 (31.25) |
| Mutation zygosity – n (%)<br>Heterozygous<br>N/A | 15 (93.75)<br>1 (6.25) |
| In-silico predicted function – n (%)<br>PTV<br>LOF | 4 (25)<br>12 (75) |
| Phenotype – n (%)<br>Dravet syndrome<br>GEFS+ | 15 (93.75)<br>1 (6.25) |
| Age at seizure onset – range in months (median) | 2 - 24 (4.25) |
| Seizure type at onset – n (%)<sup>f</sup><br>Asymmetric Tonic<br>Febrile Focal Motor<br>Febrile GTC<br>Focal Motor | 1 (6.25)<br>2 (12.5)<br>9 (56.25)<br>4 (25.0) |
| VNS related characteristics<br>Age at VNS implantation – months (median)<br>Follow-up after VNS – months (median)<br>Generator current in mA – median (range) at last follow up<br>Duty Cycle in % – median (range) at last follow-up | 21.2 – 192.7 (72.95)<br>19 – 167.5 (94.8)<br>1.75 (0.5 – 2.5)<br>21.7 (9.1 – 38.5) |
| <sup>f</sup> GTC – Generalized tonic clonic<br><sup>ø</sup> Vagus Nerve Stimulation |  |

## Results

### Patient Characteristics

Sixteen DRE patients with pathogenic or likely pathogenic variants in *SCN1A* and ≥12 months of post-VNS follow-up were included. Of these, 93.75% (15/16) were diagnosed with Dravet Syndrome while one patient had GEFS+ (Table 1). This patient had late-onset seizures (24 months), no developmental delay but persistent dialeptic spells warranting VNS implantation. The median age at seizure onset was 4.25 months (range 2-24), with a median follow-up of 141.45 months (Table 1). Most patients presented with febrile generalized or focal seizures (11/16; 68.75%) at seizure onset. The median age at VNS implantation was 72.95 months with a median post-VNS follow-up of 94.80 months (Table 1). The cohort’s demographic profile included significant underserved populations (∼70% Asian, Black and Hispanic patients; Table 1), potentially enhancing the generalizability of findings to a broader *SCN1A*-related epilepsy population. 25% (4/16) of patients were truncating variant and 75% (12/16) missense variant carriers (Table 1, Figure 1A, Table S1).

### VNS Achieves Sustained Seizure Reduction

As long-term seizure reduction in *SCN1A*-related epilepsy has not been well studied, we analyzed seizure frequency across 12-month and ten-year follow-up periods after VNS implantation to identify its long-term efficacy. We defined responders as patients exhibiting ≥50% reduction in seizure frequency at both follow-up intervals. Response was observed in 93.75% (15/16) of patients at 12 months and 87.5% (14/15) of patients at ten years (Figure 1B). Seizure frequency reduction in the 12-month (median reduction = 69.7%) and ten-year interval (median reduction = 71.6%) indicated sustained efficacy of VNS (Figure 1C). Next, we calculated Z-score normalized seizure trajectories to identify the temporal sequence of seizure frequency reduction. We observed an initial reduction followed by long-term stabilization of seizure frequency (Figures 1D-E). In summary, VNS implantation led to long-term reduction of seizures in most patients in our cohort.

### Seizure Reduction with VNS Is Associated with Higher Duty Cycles and Independent of ASM Use

Since concurrent ASM use is a significant confounder when assessing seizure frequency, particularly in DRE, we used linear mixed-effects models to quantify the confounding effect of ASM (Figures 1F). We analyzed the effect of VNS status (binary variable for pre- and post-implantation periods) and number of ASM (separated by mechanism) on seizure frequency. VNS implantation had the largest negative estimate size (-1.09), indicating it is the strongest predictor of seizure frequency reduction. As this model corrects for ASM use, this result emphasizes that VNS exerts a therapeutic effect independent of concurrent ASM use (Figure 1F).

Next, we explored whether individual VNS parameters influenced seizure reduction at the ten-year interval. Due to high correlation between the binary VNS status variable and VNS-related parameters, we analyzed the effect of VNS duration, duty cycle and generator current (mA) in a separate model (Figure 1G). Duty cycle (median = 21.7%) had the largest negative estimate size (-1.81), indicating it is a strong predictor of seizure frequency reduction. The VNS therapy duration (estimate size 0.00) was not associated with additional seizure reduction (Figure 1G), aligning with stable long-term trajectories seen in Figures 1D-E.

### Clinical course of patients in this study

All patients reported good VNS tolerance and no major serious adverse events (AEs) as shown previously^12^. Three patients experienced minor AEs, including transient hoarseness or throat discomfort, which resolved without surgical revision or discontinuation of VNS therapy (Table S1). Patient logs indicated anecdotal evidence of reduction in seizure duration after use of VNS magnets as an abortive measure. All, but one, patients are still alive at the time of this manuscript; the deceased patient experienced respiratory complications during an admission for status epilepticus.

## Discussion

Our study provides long-term data on the efficacy of VNS in patients with *SCN1A*-related epilepsy syndromes. Our findings demonstrate that VNS offers sustained seizure reduction, with most patients achieving ≥50% seizure reduction at both at both 12-month and ten-year follow-ups. The high response rate is better when compared to previous studies analyzing VNS in refractory epilepsy but is especially noteworthy in this genetically defined population^4,5^.

Importantly, our analysis showed that seizure reduction persisted after considering ASM as confounders, reinforcing VNS as a valuable adjunctive intervention. Why VNS shows efficacy in DS can only be speculated. It possibly promotes inhibitory neuronal activity in the nucleus tractus solitarius (NTS) and higher cortical regions, countering the excitation-inhibition imbalance caused by *SCN1A* variants^13^. This mechanism would align with findings that GABAergic agonists or glutamatergic antagonists in the NTS can abort seizures^14^, highlighting VNS’s potential role in creating an inhibitory environment to reduce epileptogenic activity.

We did not observe a stronger seizure frequency reduction with longer VNS use durations as one would expect based on findings in previous large VNS cohorts^15^. This effect is likely secondary to the highly refractory nature of early-onset DRE^16^. Additionally, we found that higher VNS duty cycles were correlated with improved outcomes, aligning with previously observed benefits of rapid cycling^17^. The median duty cycle in our cohort (21.7%) is comparable with the target duty cycle (17.1%) at which ideal responses have been noted previously^18^. Overall, these findings indicate that personalized VNS settings could enhance treatment efficacy. However, our data demonstrates that initiation of VNS is still delayed (median implantation ∼6 years; Table 1). Such delays may contribute to further accrual of morbidity and mortality in DRE such as DS.

While our study provides robust long-term data, limitations include the retrospective design, relatively small sample size, and patient-reported seizure data, which introduce recall bias. Variability in VNS settings across centers may also influence outcomes. Despite these limitations, our study underscores the importance of long-term follow-up and highlights the need for standardized protocols in future prospective studies. Although we cannot rule out seizure reduction is part of the normal course of Dravet Syndrome, our study demonstrates an acute 12 month after implantation response and long-term maintenance of seizure response.

## Conclusion

Our study underlines the value of genotype-driven therapeutic interventions for management of drug-resistant epilepsies associated with monogenic neurodevelopmental disorders. By demonstrating the long-term efficacy of VNS independent of ASM adjustments in *SCN1A* patients, we highlight the importance of early genetic testing to identify at-risk patients and expedite treatment options such as VNS. Future research focusing on larger, multi-center studies to validate these results and optimize the use of VNS in *SCN1A*-related epilepsy are crucial. Furthermore, these results prompt further exploration of personalized treatment strategies as well as the impact of VNS on comorbidities, such as gastrointestinal dysmotility, for refractory epilepsy patients.

## Author Contributions

S.B., A.I. and D.L. conceived the project and designed analyses.

S.B. acquired data, performed all analyses and wrote the manuscript. A.I. provided supervision and revised the manuscript. T.B., C.L., and L.M. performed variant interpretation and modeling. E.M.C.S., E.H., G.T., M.S. and R.A.T. established the database and assisted with EHR queries. M.W.W., J.L., I.K., S.P., E.P.K., and G.V.A. were primary epileptologists for the patients and M.S. was the primary neurosurgeon. S.P., S.D.L., P.K., A.A., E.P.K., G.V.A., and D.L. provided supervision and revised the manuscript. D.L. provided supervision and revised the manuscript.

## Acknowledgments

This study was supported by the National Institutes of Health (NIH) National Institute of Neurological Disorders and Stroke (NINDS) under grant R01 NS117544 (Principal Investigator: D.L.).

We thank all Lal lab members for their continued support and avid discussions. We thank other members of the Texas Institute of Restorative Neurotechnologies (TIRN) for suggestions and guidance. We thank Karol Volante for assisting with IT-related affairs.

## Data Availability Statement

The code for data analysis can be openly accessed on Github: https://github.com/Sunanjay/SCN1A_VNS/

## References

1. Gallagher D, Pérez-Palma E, Bruenger T, Ghanty I, Brilstra E, Ceulemans B, et al. Genotype–phenotype associations in 1018 individuals with SCN1A-related epilepsies. Epilepsia. 2024; 65(4):1046–59.

2. Orosz I, McCormick D, Zamponi N, Varadkar S, Feucht M, Parain D, et al. Vagus nerve stimulation for drug-resistant epilepsy: A European long-term study up to 24 months in 347 children. Epilepsia. 2014; 55(10):1576–84.

3. Samanta D, Haneef Z, Albert GW, Naik S, Reeders PC, Jain P, et al. Neuromodulation strategies in developmental and epileptic encephalopathies. Epilepsy Behav. 2024; 160:110067.

4. Dibué-Adjei M, Fischer I, Steiger H-J, Kamp MA. Efficacy of adjunctive vagus nerve stimulation in patients with Dravet syndrome: A meta-analysis of 68 patients. Seizure. 2017; 50:147–52.

5. Chen S, Li M, Huang M. Vagus nerve stimulation for the therapy of Dravet syndrome: a systematic review and meta-analysis. Front Neurol. 2024; 15:1402989.

6. Youn SE, Jung DE, Kang H-C, Kim HD. Long-term results of vagus nerve stimulation in children with Dravet syndrome: Time-dependent, delayed antiepileptic effect. Epilepsy Res. 2021; 174:106665.

7. Richards S, Aziz N, Bale S, Bick D, Das S, Gastier-Foster J, et al. Standards and guidelines for the interpretation of sequence variants: a joint consensus recommendation of the American College of Medical Genetics and Genomics and the Association for Molecular Pathology. Genet Med. 2015; 17(5):405–23.

8. Zuberi SM, Wirrell E, Yozawitz E, Wilmshurst JM, Specchio N, Riney K, et al. ILAE classification and definition of epilepsy syndromes with onset in neonates and infants: Position statement by the ILAE Task Force on Nosology and Definitions. Epilepsia. 2022; 63(6):1349–97.

9. Brunklaus A, Ellis R, Reavey E, Forbes GH, Zuberi SM. Prognostic, clinical and demographic features in SCN1A mutation-positive Dravet syndrome. Brain. 2012; 135(8):2329–36.

10. Wirrell EC, Laux L, Donner E, Jette N, Knupp K, Meskis MA, et al. Optimizing the Diagnosis and Management of Dravet Syndrome: Recommendations From a North American Consensus Panel. Pediatr Neurol. 2017; 68:18–34.e3.

11. Blume WT, Lüders HO, Mizrahi E, Tassinari C, Boas WVE, Engel J. Glossary of Descriptive Terminology for Ictal Semiology: Report of the ILAE Task Force on Classification and Terminology. Epilepsia. 2001; 42(9):1212–8.

12. Sirsi D, Khan M, Arnold S. Vagal Nerve Stimulation: Is It Effective in Children with Dravet Syndrome? J Pediatr Epilepsy. 2015; 05(01):007–10.

13. Rutecki P. Anatomical, Physiological, and Theoretical Basis for the Antiepileptic Effect of Vagus Nerve Stimulation. Epilepsia. 1990; 31(s2):S1–6.

14. Walker BR, Easton A, Gale K. Regulation of Limbic Motor Seizures by GABA and Glutamate Transmission in Nucleus Tractus Solitarius. Epilepsia. 1999; 40(8):1051–7.

15. Elliott RE, Morsi A, Tanweer O, Grobelny B, Geller E, Carlson C, et al. Efficacy of vagus nerve stimulation over time: Review of 65 consecutive patients with treatment-resistant epilepsy treated with VNS >10years. Epilepsy Behav. 2011; 20(3):478–83.

16. Tamura K, Sasaki R, Sakakibara T, Dahal R, Takeshima Y, Matsuda R, et al. Additional Effect of High-output Current and/or High-duty Cycle in Vagus Nerve Stimulation for Adolescent/Adult Intractable Epilepsy. Neurol medico-Chir. 2023; 63(7):273–82.

17. Kayyali H, Abdelmoity S, Bansal L, Kaufman C, Smith K, Fecske E, et al. The Efficacy and Safety of Rapid Cycling Vagus Nerve Stimulation in Children With Intractable Epilepsy. Pediatr Neurol. 2020; 109:35–8.

18. Fahoum F, Boffini M, Kann L, Faini S, Gordon C, Tzadok M, et al. VNS parameters for clinical response in Epilepsy. Brain Stimul. 2022; 15(3):814–21.

